# High-Throughput Genotyping of *Plasmodium vivax* in the Peruvian Amazon via Molecular Inversion Probes

**DOI:** 10.1101/2024.06.27.24309599

**Authors:** Zachary R. Popkin-Hall, Karamoko Niaré, Rebecca Crudale, Alfred Simkin, Abebe A. Fola, Juan F. Sanchez, Danielle L. Pannebaker, David J. Giesbrecht, Isaac E. Kim, Özkan Aydemir, Jeffrey A. Bailey, Hugo O. Valdivia, Jonathan J. Juliano

## Abstract

*Plasmodium vivax* transmission occurs throughout the tropics and is an emerging threat in areas of *Plasmodium falciparum* decline, causing relapse infections that complicate treatment and control. Targeted sequencing for *P. falciparum* has been widely deployed to detect population structure and the geographic spread of antimalarial and diagnostic resistance. However, there are fewer such tools for *P. vivax*. Leveraging global variation data, we designed four molecular inversion probe (MIP) genotyping panels targeting geographically differentiating SNPs, neutral SNPs, putative antimalarial resistance genes, and vaccine candidate genes. We deployed these MIP panels on 866 infections from the Peruvian Amazon and identified transmission networks with clonality (IBD>0.99), copy number variation in *Pvdbp* and multiple *Pvrbps*, fixation of putative antimalarial resistance, and balancing selection in 13 vaccine candidate genes. Our MIP panels are the broadest genotyping panel currently available and are poised for successful deployment in other regions of *P. vivax* transmission.

## Introduction

*Plasmodium vivax* is the most geographically widespread malaria species, with active transmission throughout South and Southeast Asia, Latin America, the Pacific, and parts of Africa^1^. *P. vivax* is also an emerging threat in areas where *P. falciparum* has been eliminated or is on track for elimination^2^. Transmission of *P. vivax* is more complex due to the presence of hypnozoites in the liver, which cause disease relapse without a new infectious mosquito bite^3^, and earlier gametocytogenesis than *P. falciparum*^4^, meaning that it can be transmitted earlier in an infection, even among asymptomatic patients^5^. Genomic epidemiology is a powerful tool for understanding parasite biology and transmission, but most work has been done in *P. falciparum*^6–11^. Given the challenging biology impeding control^12^, there is an urgent need for the development of high-throughput genotyping methods to better understand this neglected parasite.

Genomic epidemiology studies of *P. falciparum* using whole genome sequencing (WGS), molecular inversion probes (MIPs), and amplicon sequencing have generated useful data to support malaria elimination by identifying parasite population structure^6,7^, importation and transmission networks and connectivity^7^, and the geographical spread of antimalarial^6,7,9–11,13–15^ and diagnostic resistance^16^. Such studies of *P. vivax* are rarer but similarly necessary, especially in South America, where *P. vivax* is now the predominant species and sympatric with *P. falciparum*^17^. Peru had the fourth largest number of malaria cases in the Americas in 2022, with the majority being *P. vivax*^18^. Most transmission occurs in the Amazonian region and department of Loreto^19,20^, where morbidity and economic losses stemming from malaria are greater than in the rest of the country^21^. Cases tend to cluster at the household and community levels^22^, with most malaria cases being asymptomatic and submicroscopic^22–24^. While the malaria burden in Peru has significantly declined from 1990 levels, progress has stalled and even reversed in the past decade^21^, with increases from 2010 onwards reversing the successes of the early 2000s^20^. Furthermore, *P. vivax* prevalence has been slower to decline than *P. falciparum* due to its unique biology that makes it more resistant to most current control measures, which are designed to target *P. falciparum*^25^.

In contrast to *P. falciparum*, where there are multiple large-scale genotyping panels in use ^6,7,9–11,13–16^, there are relatively few existing panels for *P. vivax*. Genotyping panels based on targeted deep sequencing have been used to study *P. falciparum* populations and their capacity to escape control interventions in Africa and Latin America, including population structure and gene flow, geographic transmission patterns, origins of imported malaria cases, spread of antimalarial drug resistance, and genetic diversion of vaccine targets. ^6,7,9–11,13–16^. *P. vivax* genotyping, however, has historically relied on microsatellite genotyping with SNP barcodes and multiplex amplicon sequencing panels only more recently being used in focal studies ^26–31^. A summary of existing panels is shown in **Table 1**.

**Table 1.** Summary of Existing *P. vivax* Genotyping Panels.

| Method | Targets Included | Neutral Sites | Antimalarial Resistance | Vaccine Candidates | Advantages | Disadvantages | Reference |
| --- | --- | --- | --- | --- | --- | --- | --- |
| High resolution melting (HRM) SNP barcode | 42 SNPs | Yes | No | No | Economical, sequencing not required, effectively differentiates samples by geography to characterize population genomics and identify importation, requires minimal sample volumes and concentrations | Does not incorporate information on biologically significant genes (antimalarial resistance or cell invasion ligands), limited ability to detect selection or population dynamics | 27 |
| Papua New Guinea SNP barcode | 146 SNPs | Yes | No | No | Reveals subnational population structure by geography, allows detection of within-country transmission networks, can detect polyclonal infections | Does not incorporate information on biologically significant genes (antimalarial resistance or cell invasion ligands) | 29 |
| vivaxGEN-geo | 33, 50, or 55 SNPs (three options available) | No | Limited | No | Accurately identifies importation on a global level, reflects global population diversity, easily modifiable, works with polyclonal infections | Non-neutral SNPs may need to be replaced over time, limited South Asian data incorporated, does not systematically address biologically significant genes (antimalarial resistance or cell invasion ligands) | 28 |
| Pv AmpliSeq | 229 amplicons (includes 33 vivaxGEN-geo SNPs and 42-SNP Vietnam barcode, 11 putative drug resistance genes) | No | Yes | Limited to *Pvama1* | Provides high resolution within a high-transmission country with high gene flow (Vietnam), incorporates vivaxGEN-geo to identify importation, includes 11 putative antimalarial resistance genes, can be used to calculate identity-by-descent (IBD), works with low parasitemia (5 parasites/μL) samples, incorporates *Pvama1* to facilitate study of polyclonal infections and antigen diversity, easily modifiable | Needs to be updated for new geographic contexts, does not include the full suite of putative antimalarial resistance genes, unable to detect selection in cell invasion ligands, identification of importation origin not always successful | 26 |
| Pv AmpliSeq v2 | 33 vivaxGEN-geo SNPs, 41-SNP Peru barcode, 11 putative drug resistance genes | No | Yes | Limited to *Pvama1* | Builds on Pv AmpliSeq v1 to characterize Peruvian population structure, incorporates the core SNPs and associated advantages from PvAmpliSeq v1, likely to work elsewhere in Latin America | Needs to be updated for new geographic contexts, does not include the full suite of putative antimalarial resistance genes, unable to detect selection in cell invasion ligands, identification of importation origin not always successful | 30 |
| Microhaplotype<br>Panels | 100<br>microhaplotypes<br>(3-10 SNPs per<br>each) | Yes | No | No | Works reliably on paired samples<br>to distinguish relapse and<br>recrudescence from reinfection,<br>incorporates global diversity for a<br>standardized barcode that can be<br>used anywhere and enable the<br>detection of importation, enables<br>high-resolution population<br>structure analysis based on IBD,<br>can detect polyclonal infections | Markers were<br>selected with<br>relatively small<br>number of<br>samples from<br>South Asia and<br>Latin America,<br>does not<br>incorporate<br>putative vaccine<br>candidates or<br>antimalarial<br>resistance genes | 31 |

MIPs are a highly multiplexed targeted sequencing approach that allow thousands of loci to be captured in a single tube. They are high-throughput and cost-effective for large genomic studies^32^ and have several advantages including the ability to tile across genes and integrated unique molecular identifiers (UMIs) that tag specific strands of DNA and allow bioinformatic error correction. This approach has been highly successfully used for studying *P. falciparum* epidemiology in Africa ^6,7,9,10^. Thus, we sought to replicate it by creating four panels of MIPs to address multiple pertinent questions regarding *P. vivax* biology, including population structure, putative antimalarial resistance, and selection in potential blood-stage vaccine targets. Using publicly available *P. vivax* WGS datasets (details in **Supplemental Material**), we designed our panels based on all known *P. vivax* genetic diversity to make them deployable in any epidemiological context. Here, we report the use of these MIP panels on 866 *P. vivax* samples collected in the Peruvian Amazon by NAMRU SOUTH between 2011 and 2018 which enabled us to detect the frequency of putative antimalarial resistance alleles, identify balancing selection on red blood cell (RBC) invasion ligands, calculate complexity of infection, detect copy number variation in multiple genes, and reveal highly related (IBD ≥ 0.99) clonal transmission networks that span years.

## Results

High-Throughput Sequencing and Genotyping of Peruvian *P. vivax* Isolates

Following panel optimization and MIP rebalancing on high parasitemia isolates, we performed MIP captures on 866 samples collected by NAMRU South, primarily in Loreto (**Figure 1**). Two captures were performed for each sample, with one for the tiled gene panel (PvG) and one multiplexed capture for three SNP panels (PvFST, PvMAF5, PvMAF40), with a repool (only samples with average MIP <100X UMI coverage included in repool) after initial sequencing to improve sequencing coverage and depth. We successfully extracted usable sequences from 685 samples. Additional details on sequencing output (**Supplemental Figure 1**), samples successfully sequenced (**Supplemental Table 1**) and impact of parasitemia (**Supplemental Figure 2**) are described in the **Supplemental Material**. The number of samples retained for each step of the analysis are shown in **Supplemental Figure 3**.

**Figure 1.**
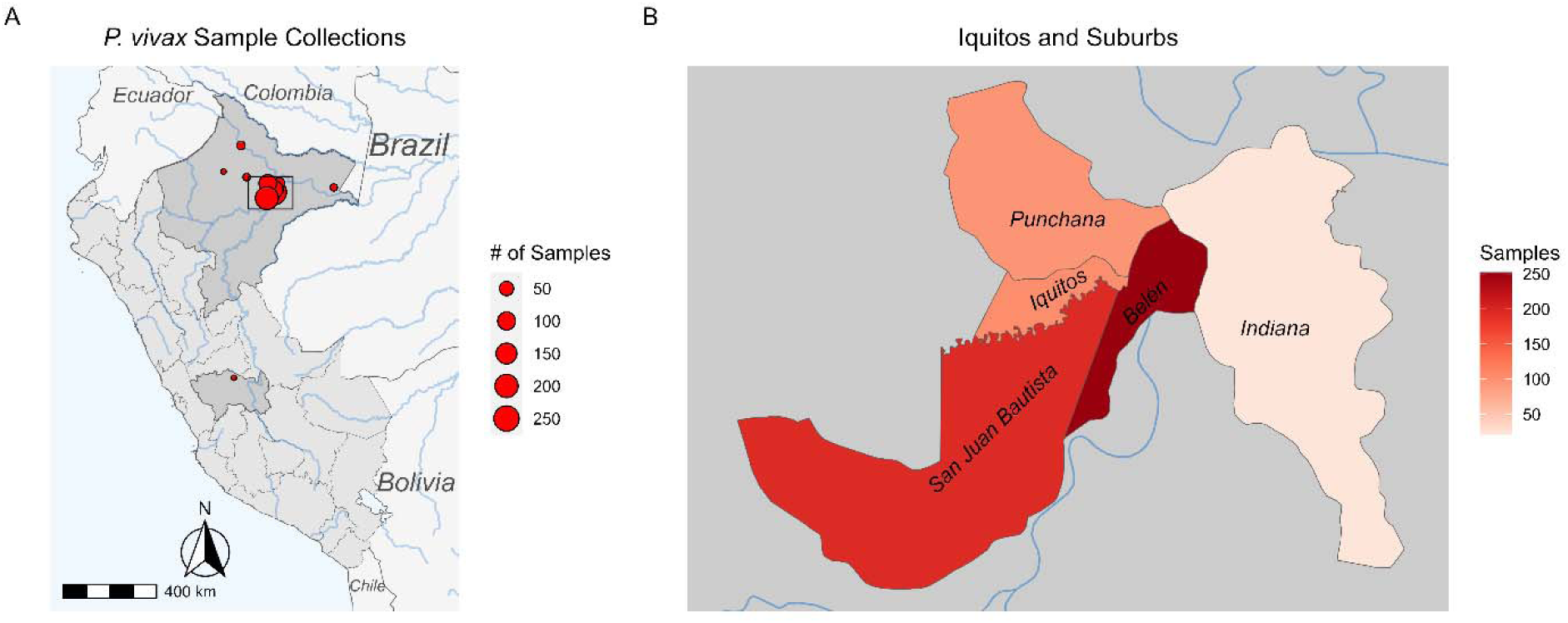
Study sites and sample size. **A)** Locations of sample collection by community within Peru, scaled by size of circle. Darker gray boundaries identify the regions and departments of Loreto (where all but one of the circles are located) and Junín. Blue rectangle defines Iquitos, expanded in **B)** Inset map depicting region around Iquitos, scaled by color of shading. Map created in R using open data from GADM database, v 3.6. www.gadm.org and the Sf package (https://cran.r-project.org/package=sf) with GPL-2 license (https://cran.r-project.org/web/licenses/GPL-2)

### Clonal Outbreak Detected in Belén 2012-2014

To assess relatedness between isolates, a principal component analysis (PCA) was performed for 685 samples that passed quality filters and had sufficient associated metadata for further analysis. The PCA revealed no broad geographical or temporal structure, but PC1 and PC2 included multiple clusters that were associated by both city and year (**Supplemental Figure 4**). To investigate these clusters further, pairwise identity by descent (IBD) was calculated for these samples using the MIPanalyzer R package (https://github.com/mrc-ide/MIPanalyzer). The median, mean, and maximum pairwise IBD were 0, 0.0878, 0.99, respectively. The overall distribution was bimodal, with 599 isolates being distantly related to at least one other pair (IBD < 0.25) and a 448 being very highly related to at least one other pair (IBD ≥ 0.9) (**Supplemental Figure 5**). To detect evidence of clonal transmission, IBD networks were generated between isolates with IBD ≥ 0.99 for five communities with a sufficient number of isolates: Belén, Indiana, Iquitos, Punchana, and San Juan Bautista (**Figure 2**). In Belén, multiple clonal clusters were detected (**Figure 2**), with many of these clusters spanning multiple years of collection. Smaller clonal networks were also detected in isolates collected from 2016 and 2017 in Indiana, 2012 and 2013 in Punchana, 2016 and 2017 in Punchana, and 2015-2017 in San Juan Bautista (**Figure 2**).

**Figure 2.**
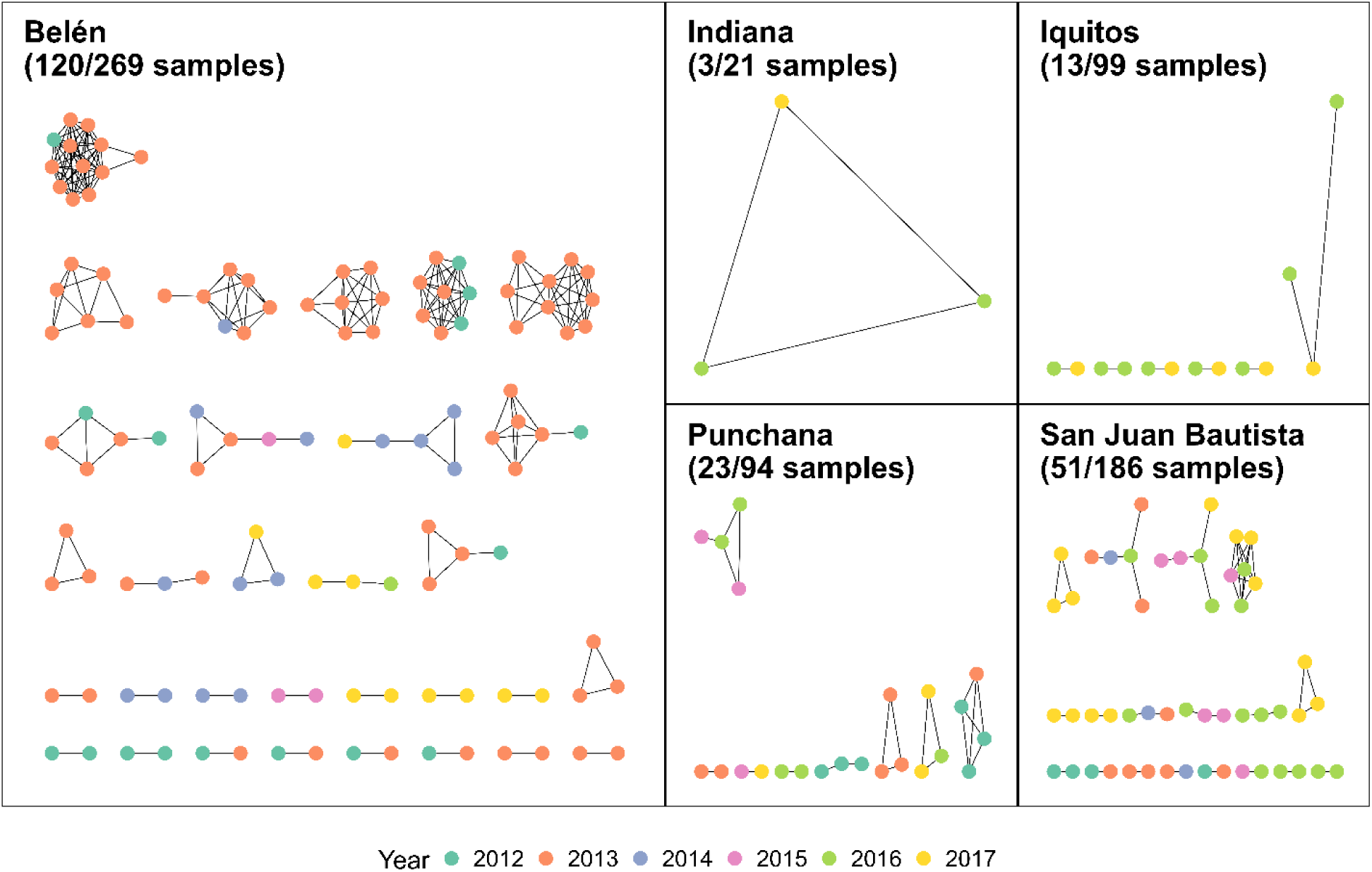
Identity by descent (IBD) networks for five communities within the vicinity of Iquitos: Belén, Indiana, Iquitos, Punchana, and San Juan Bautista. Each network node is a sample, with the edges representing IBD estimates. Nodes are color coded by collection year. An IBD threshold of 0.99 was used to generate networks. The Belén network shows evidence of a clonal outbreak in 2013 that is closely related to isolates collected in 2012 and 2014. In all five cities, there are networks connecting closely related isolates across years.

### Most Infections Are Monoclonal

Of 697 isolates (all samples that passed quality filters, regardless of metadata) analyzed with THEREALMcCOIL, the majority (637, 91.4%) were monoclonal (**Figure 3**). Among the 60 isolates estimated to be polyclonal, 44 (73.3%) were estimated to contain two clones, 14 (23.3%) were estimated to contain three, and two (3.3%) were estimated to contain four clones. The median COI estimate across all samples was 1.

**Figure 3.**
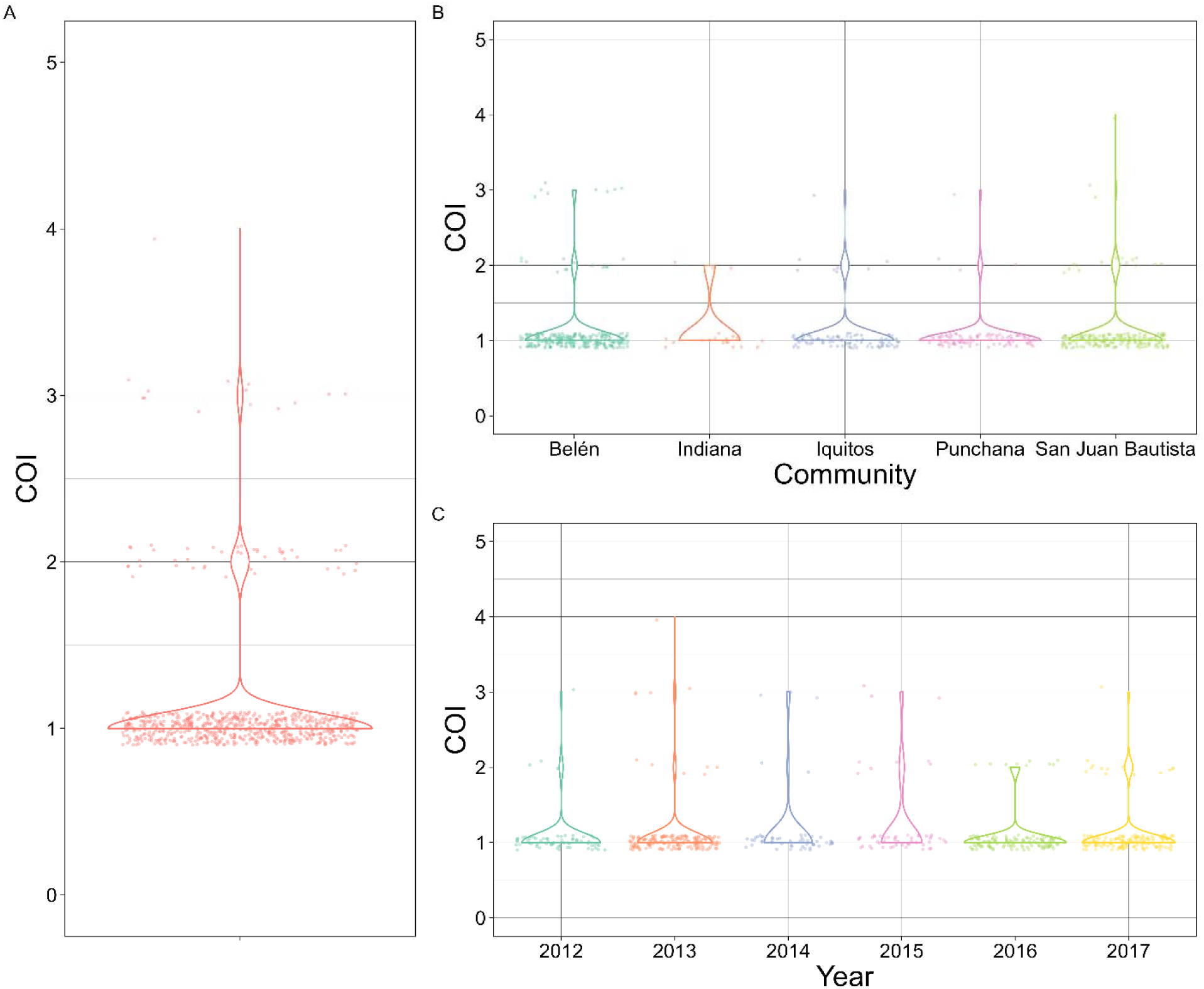
Violin plot depicting complexity of infection (COI) of: **A) all *P. vivax* isolates analyzed.** Each dot represents the mean COI estimate for a given isolate. The majority of isolates (91.4%, n = 637/697) are monoclonal, with 44 isolates (6.31%) containing two clones,14 isolates (2.01%) containing three clones, and two isolates (0.287%) containing four clones. **B) *P. vivax* isolates from five communities with largest sample numbers**. COI does not differ significantly by community. **C) *P. vivax* isolates from the same five communities ordered by collection year**. COI does not differ significantly by year.

### Copy Number Variation Detected in Duffy and Reticulocyte Binding Proteins

Prior to calculating copy number based on UMI depth, the larger dataset of 685 samples was filtered for missingness and high coverage (see Methods for details), as well as monoclonality, leaving 595 samples for copy number calculation. Copy number variants (CNV) were detected in twelve samples (2.02%) (**Figure 4**). All twelve samples contained multiple copies of reticulocyte binding protein 2b (*Pvrbp2b*), eleven of which had two copies while one had three copies. Ten of the twelve samples showed CNV only in *Pvrbp2b*, while two had additional duplicated genes (**Figure 4**). The sample with three copies of *Pvrbp2b* also had two copies of Duffy Binding Protein (*Pvdbp*), *Pvrbp1b*, *Pvrbp2a,* and *Pvrbp2c* (**Figure 4**). The final sample had a duplication in *Pvrbp2c*. No CNV were detected in either *Pvrbp1a* or erythrocyte binding protein (*Pvebp*).

**Figure 4.**
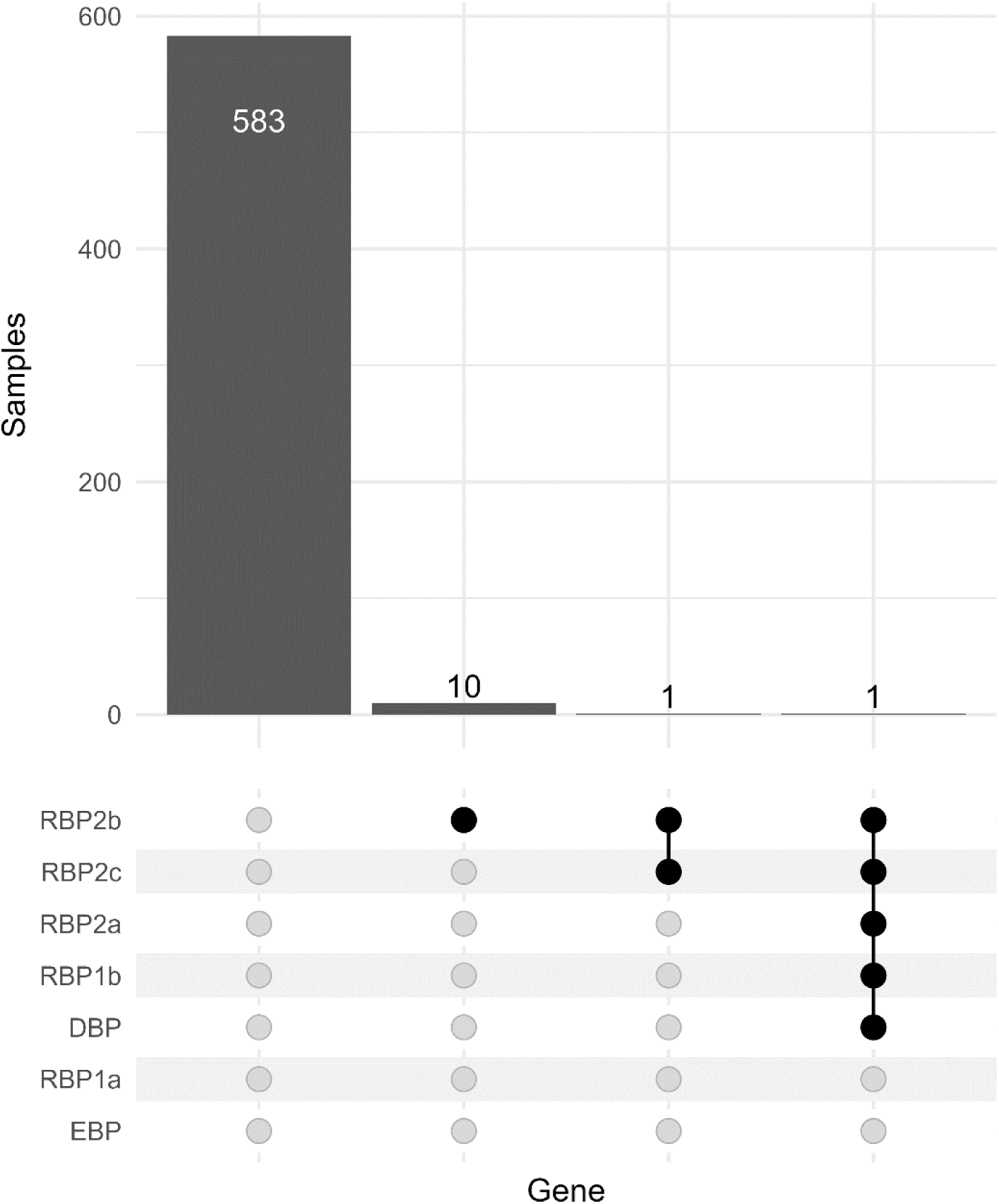
Upset plot showing the frequency of copy number variation (CNV) among monoclonal isolates. The majority of isolates (98.0%) do not exhibit CNV. CNV in *Pvrbp2b* was observed in twelve (2.01%) samples, with one isolate having three copies, as well as duplications in *Pvdbp*, *Pvrbp1b*, *Pvrbp2a*, and *Pvrbp2c*. One other isolate showed duplications in both *Pvrbp2b* and *Pvrbp2c*. No CNV was detected in either *Pvebp* or *Pvrbp1a*.

### Putative Antimalarial Resistance Mutations at or Approaching Fixation

Allele frequencies were calculated for mutations that are putatively involved in antimalarial resistance in *P. vivax*. The full set of allele frequencies for all putative antimalarial resistance genes is reported in **Supplemental Table 3**. Mutations were detected in *Pvcrt*, *Pvdhfr-ts*, *Pvpppk-dhps*, and *Pvmdr1* (**Table 1**). Allele frequencies in detected mutations vary from rare (1.6%) to complete fixation (100%) (**Table 1**). In addition to the *Pvmdr1* S698G mutation, which is completely fixed, two additional mutations in *Pvmdr1* are approaching fixation: F976Y (95%) and L1076F (91.3%) (**Table 1**). The *Pvpppk-dhps* G383A mutation was also detected at a relatively high frequency (49.6%), while all other detected mutations were detected at frequencies below 15% (**Table 1**).

**Table 1.** Frequencies of known putative drug resistance mutations detected in sample set. Three alleles in *Pvmdr1* are at or approaching fixation.

| **Gene** | **Mutation** | **Drug(s)** | **Study Frequency** | **Global Frequency<sup>33</sup>** |
| --- | --- | --- | --- | --- |
| CRT | K9 duplication | CQ | 11.0% | 23.9% |
| DHFR | F57L | SP, AQ | 1.6% | 5.5% |
| DHFR | N50I | SP | 0.0% | 0.7% |
| DHFR | S58R | SP, AQ | 0.0% | 6.2% |
| DHFR | N117S | SP, AQ | 3.3% | 8.4% |
| DHFR | I173L | SP | 5.7% | 1.1% |
| DHPS | G383A | SP | 49.6% | 29.3% |
| DHPS | 553G | SP | 0.0% | 17.8% |
| MDR1 | S513R | CQ | 0.0% | 18.2% |
| MDR1 | S698G | CQ | 100.0% | 22.9% |
| MDR1 | L908M | CQ | 7.9% | 2.2% |
| MDR1 | M958T | CQ | 1.9% | 0.5% |
| MDR1 | F976Y | CQ, SP, AQ | 95.0% | 48.9% |
| MDR1 | L1076F | CQ, SP, AQ | 91.3% | 22.0% |

### Potential Balancing Selection in Multiple RBC Invasion Ligands

In addition to determining the allele frequencies of putative antimalarial resistance markers, we also calculated Tajima’s D in 100bp bins with a 50bp step size across loci encoding merozoite surface and secreted proteins that are involved in the red blood cell invasion that we tiled with the PvG MIP panel. These ligands are potential *P. vivax* vaccine targets (see **Supplemental Material)**. The strongest signatures of balancing selection (Tajima’s D > 2), were detected in 13 of 20 genes analyzed, including apical membrane antigen 1 (*Pvama1*), *Pvdbp*, *Pvebp*, merozoite surface protein 1 (*Pvmsp1*), *Pvrbp1a*, *Pvrbp1b*, *Pvrbp2a*, *Pvrbp2b*, *Pvrbp2c*, tryptophan-rich antigen 111 (*Pvtrag11*), *Pvtrag19*, *Pvtrag22*, *Pvtrag34*, and *Pvtrag36* (**Figure 5**). The median value across all RBC invasion ligands was -0.577, while the minimum was -1.39 (*Pvrbp1a*) and the maximum was 3.14 (*Pvtrag11*). No Tajima’s D values <-2, consistent with strong directional selection, were detected in any RBC invasion ligand. All polymorphisms detected in RBC invasion ligands are reported in **Supplemental Table 4** and all Tajima’s D values are shown in **Supplemental Figure 6**.

**Figure 5.**
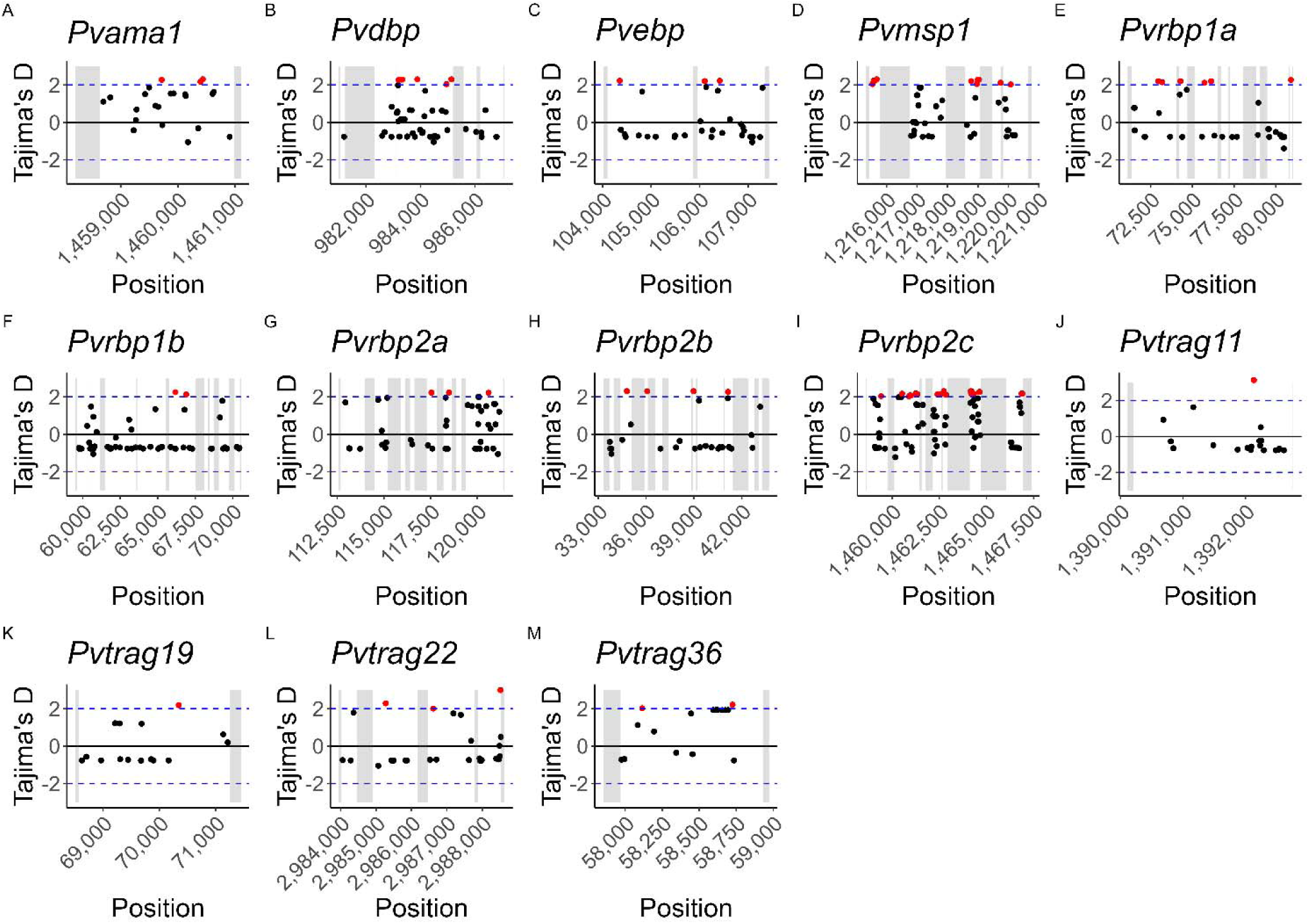
Scatter plots depicting Tajima’s D values in 100bp bins with a 50bp step size across potential vaccine target genes. Grey boxes indicate gene regions that were not captured by MIPs. Only genes with Tajima’s D values >2, i.e. showing strong signatures of balancing selection, are depicted. Dots are placed in the midpoint of each bin, with values >|2| highlighted in red.

## Discussion

In this study, we demonstrate that the high-throughput MIP genotyping approach used to characterize the genomic epidemiology of *P. falciparum* in Africa is also effective and informative for South American *P. vivax* parasite populations. Combined, our novel genotyping panels are the broadest genotyping panel currently available, working reliably on *P. vivax* isolates with parasitemias ≥100 parasites/µL to characterize population structure, relatedness, putative antimalarial resistance mutation frequency, and identify signatures of selection in candidate vaccine targets. By deploying them on samples from Peru, we were able to simultaneously detect highly related (IBD ≥ 0.99) samples within the same community across years, detect CNV in *Pvdbp* and multiple *Pvrbps*, estimate COI, identify multiple putative antimalarial resistance genes at or near fixation, and detect balancing selection in 13 vaccine candidate genes.

These new MIP panels build on both the success of *P. falciparum* MIPs and other existing *P. vivax* genotyping panels to enable a thorough analysis of multiple key elements of *P. vivax* biology. Crucially, like the microhaplotype panels^31^ and unlike the existing amplicon panels^26,30^, our MIP panels should be deployable in any geographic context without modifications. In addition, unlike any existing panels, our MIP panels contain both geographically discriminating and neutral SNPs in addition to 21 vaccine candidate genes and five genes that are putatively associated with drug resistance. This single set of panels therefore enables the simultaneous detection of population structure and transmission networks along with surveillance of potential variants of concern for antimalarial resistance and mutations that may decrease the effectiveness of vaccines.

While our MIP panels are efficient, reliable, and high-throughput, they nevertheless will require further refinement to better meet every need for a widely deployed *P. vivax* genotyping panel. As with the amplicon, SNP, and microhaplotype panels, MIPs require sophisticated molecular laboratories with sequencing capabilities, which are rare in malaria-endemic regions. In addition, the computational analyses necessary to analyze the genotyping results require substantial bioinformatic resources and expertise, which are also in short supply in malaria-endemic regions. Another limitation of both the MIP and amplicon panels is that the incorporation of putative antimalarial resistance mutations is of questionable significance to malaria control programs since none of these mutations are validated. In addition, unlike the microhaplotype panels, the MIP panels have not been validated for use in treatment efficacy studies (TES), which are critical to distinguish reinfection from recrudescence and relapse. Finally, a major limitation of MIPs is inconsistent performance in low-parasitemia samples, which hampers highly-multiplex amplicon panels as well. Since *P. vivax* typically causes low-density infections, many of which are undiagnosed in the Amazon^24^, we are currently limited to analyzing the high-density minority of infections, which could mean missing vital population data.

In addition to demonstrating the viability of MIPs to characterize and surveil *P. vivax* in a highly endemic region, we also identified multiple aspects of Peruvian *P. vivax* biology with potential implications for malaria control and elimination plans in the country. Within the immediate vicinity of Iquitos, we identified multiple clonal (IBD ≥ 0.99) networks with temporal-geographic connections. The Amazon region is known for its high clonality^30^, which contributes to a high proportion of submicroscopic infections ^22^. Most notably in our study, the community of Belén appears to have had a clonal *P. vivax* outbreak from 2012-2014, which has also been observed elsewhere in the Peruvian Amazon^30^. Iquitos is known to be a source of *P. vivax* transmissions in the region, due to the highly mobile population^34,35^. The presence of a clonal outbreak in the suburb of Belén suggests that this strain likely spread elsewhere as well, meaning that monitoring transmission in the Iquitos region should provide a representative picture of most *P. vivax* within the Peruvian Amazon.

Several genomic epidemiology studies of *P. vivax* have occurred in Loreto with variable findings. Transmission patterns are complex, with a heterogenous spatio-temporal malaria distribution that is correlated with human mobility patterns, with travel history, and occupation influencing prevalence^22,36^. Urban areas have a similar proportion of monoclonal and polyclonal infections, but monoclonal infections predominate in rural areas and complexity of infection (COI) is higher in urban areas^37^, although most isolates regardless of urbanization are monoclonal^25,30,36,38,39^. While heterozygosity (He) is similar in all localities, multiple studies have identified substantial differentiation by geography^30,36,38^, which is also reflected in principal component analysis that separates rural and urban isolates^37^. These studies have also identified substantial substructure and clonal propagation in the parasite population^40^, with Iquitos functioning as the main source of parasites for the rest of the region^37^. Clonal populations reflected by isolates from the same spatio-temporal context clustering with high identity by descent (IBD) have been identified^40^. However, a study of isolates collected between 2015 and 2019 identified three subpopulations but no geographic substructure^25^.

CNV was rare in these samples and most frequently found in *Pvrbp2b*, although one isolate exhibited CNV in five genes. Although CNV has been previously observed in *Pvebp* ^41^, we did not detect any. CNV has previously been identified in *Pvrbp2a* and *Pvrbp2b*, with duplications detected in two Thai isolates^42^. However, duplications in *Pvrbp1b* and *Pvrbp2c* have not been previously described. The *Pvrbp* gene family plays an important role in invading reticulocytes^43–45^, is immunogenic, and is a potential vaccine target. While there are multiple functional *Pvrbp* genes, the family also contains pseudogenes^43^. The *Pvrbp*s vary in their degree of polymorphism, e.g. *Pvrbp2c* is more polymorphic than *Pvrbp1*^42^, and their different domains may be under differential selective pressure^42^. Previous analyses have not identified deviations from neutrality in the *Pvrbp*s^46,47^.

We also detected a duplication in *Pvdbp* in one isolate. *Pvdbp* plays a critical role in red blood cell (RBC) invasion in Duffy antigen/receptor for chemokines (DARC)-positive individuals, who make up the vast majority of people living in areas with high *P. vivax* transmission^48^. As such, PvDBP is a promising blood-stage vaccine candidate^48^. However, studies have demonstrated low *Pvdbp* immunogenicity in the Amazon region, and *Pvdbp* is highly polymorphic^48^. *Pvdbp* copy number variation (CNV) has been detected globally in isolates from both DARC-positive and DARC-negative individuals^49,50^. The implications of the CNV we detected in 14 isolates are somewhat unclear, but may suggest that vaccines targeting DBP or the RBPs that do not account for this variability may have reduced efficacy^51^.

While none of the putative antimalarial resistance mutations in *P. vivax* are validated, they have been associated with resistance to chloroquine (CQ)^52,53^, sulfadoxine-pyrimethamine (SP)^54–56^, and other drugs^57,58^. Since CQ is still the first-line treatment for *P. vivax* in Peru and many other contexts^18^, monitoring putatively resistance-conferring mutations in this gene is critical to maintaining efficacious treatment programs. Previous studies have identified mutations in chloroquine resistance transporter (*Pvcrt*), which have been associated with vivax CQ resistance^49^, that were nearly at fixation^36^. However, since these mutations are in intronic regions, the functional significance is unclear^36,52^. Polymorphisms in multidrug resistance protein 1 (*Pvmdr1*) have been detected at heterogeneous levels throughout the Peruvian Amazon^53^, including in the CQ resistance-associated Y976F and F1076L mutations^40,53^. In our analysis, we find the *Pvcrt* K9 duplication at a relatively low level (11%), while both *Pvmdr1* Y976F and F1076L are approaching fixation (>90%), substantially higher than the 15.2% frequency found with amplicon sequencing elsewhere in Loreto^30^. As has been found elsewhere^30,58,59^, the G698S mutation was at fixation.

SP is used as a partner drug in artemisinin combination therapy (ACT) for *P. falciparum*. In addition, while SP is not a preferred treatment for *P. vivax*, it is sometimes given in areas where *P. falciparum* and *P. vivax* co-occur, although it was removed as a first-line treatment in Peru in 2001^60^. As with *Pvcrt*, previous studies have identified mutations in dihydrofolate reductase (*Pvdhfr*) and dihydropteroate synthase (*Pvdhps*) ranging in frequency from very low (1.3%) to fixation in Peru^25,36,40^, which may have occurred while SP was still used for *P. falciparum* treatment in Peru^25^. The frequency of the *Pvdhps* A383G mutation is similar in our analysis (49.6%) to that found with amplicon sequencing (61.7%)^30^. We also found similar frequencies of the *Pvdhfr* S117N mutation (3.3% vs. 1.3%), but identified no isolates with the S58R mutation, in contrast to the relatively high frequency (54.3%) found in the other study^30^. Conversely, we identified the F57L mutation at a low frequency (1.6%), while the other study did not find it at all ^30^. Our findings also contrast with data from PNG that found the F57L and S58R mutations to be consistently linked^61^. Although the clinical and operational significance of these mutations associated with reduced CQ and SP susceptibility is unclear, monitoring them may enable Peru’s malaria elimination goals, e.g. by increasing surveillance intensity in regions where their frequency is rising.

We also demonstrate evidence of balancing selection in 13 RBC invasion ligands which are potential vaccine targets, including *Pvama1*, *Pvdbp*, *Pvebp*, *Pvmsp1*, four members of the *Pvrbp* family, and four members of the *Pvtrag* family. The biological aspects of balancing selection in these targets are discussed in the **Supplemental Material**. While the PvG panel identified potential balancing selection in 13 genes, it is limited by our ability to successfully design MIPs to target the whole genes. For some genes, such as *Pvama1*, we were unable to design MIPs targeting the beginning of the gene, while for others, such as *Pvrbp2a*, there are multiple smaller regions within the gene that we were unable to target. Other genes, such as *Pvcsp*, contain repeats that are difficult to target with the short length of the capture design. A future redesign of the PvG panel may improve coverage within these genes of interest. Nonetheless, even with this limitation, we were able to find signatures of selection.

Our four MIP panels, along with existing amplicon and microhaplotype panels, are poised to begin replicating the successful genomic epidemiology studies of *P. falciparum* in *P. vivax*. While their performance is less reliable with lower parasitemia (a common feature of *P. vivax* infections), this shortcoming may be overcome by incorporating enrichment methods such as selective whole genome amplification (sWGA) prior to MIP capture. In this analysis of Peruvian isolates, we demonstrate their utility to simultaneously detect population structure, CNV, COI, putative antimalarial resistance mutation frequencies, and signatures of selection. As malaria control programs move toward the elimination of *P. falciparum*, such a tool for *P. vivax* will be critical if indeed it expands to fill the niche left by *P. falciparum*. Future MIP analyses of isolates from other geographical regions and/or collection years will likely enable a robust characterization of global and/or temporal trends in *P. vivax* biology, and refinement of the panel for low density infections..

## Methods

### Sample Collection

Samples were collected by NAMRU SOUTH between 2011 and 2018, primarily in the vicinity of Iquitos, the major city in the Peruvian Amazon, within Loreto Department. The sampling scheme has been described in detail elsewhere^62,63^.

### Sample Processing and Sequencing

DBS were extracted using the Chelex method, as previously described^32,64^. Extracted DNA was used for molecular inversion probe (MIP) captures, which were sequenced as previously described for *P. falciparum*^7,32^. Four new MIP panels were designed for *P. vivax* based on the PvP01 genome assembly: 1) PvG, a tiled gene panel designed to densely genotype genes of interest, including putative drug resistance markers and invasion ligands that are potential vaccine targets; 2) PvFST, a panel designed to capture geographically discriminating SNPs between and within continents; 3) PvMAF5, a panel designed to capture rare neutral (non-coding) SNPs (minor allele frequency <0.05); and 4) PvMAF40, a panel designed to capture common neutral SNPs (MAF > 0.4). The details of the MIP design scheme are given in the **Supplemental Material**. The generated libraries were sequenced on an Illumina Nextseq 550 platform using 150 bp paired end sequencing with dual indexing using Nextseq 500/550 Mid-output Kit v2 in the case of the gene panels and 75 bp paired end sequencing in the case of the SNP panels.

### MIP Variant Calling and Filtering

MIP sequencing data was processed using *MIPWrangler* within *MIPTools* (https://github.com/bailey-lab/MIPTools), which normalizes read counts based on unique molecular identifiers (UMIs) that are incorporated in the MIPs (Aydemir, unpublished). Variant calling for all panels was performed using *FreeBayes*^65^ within *MIPTools*. For the SNP panels, we specified a minimum UMI depth of 3, a within-sample allele frequency threshold of 0.5, and a minimum alternate read count of 2 to obtain 616 variant SNPs (PvFST), 437 variant SNPs (PvMAF5), and 377 variant SNPs (PvMAF40). *Bcftools*^66^ v1.9 was used to filter the resulting VCF to include only targeted SNPs. The filtered VCFs were imported into R using vcfR^67^ (version 1.14), where they were assessed for sample and variant missingness. Samples with >50% missingness on any panel were excluded from the VCFs, which were then sorted and concatenated with *bcftools*. Finally, *bcftools* was used to subset the resulting VCF to include only biallelic SNPs and exclude SNPs with >50% missingness The tiled gene panel was processed in a similar fashion, yielding 13,927 variant SNPs.

### Population Structure Analysis

*MIPanalyzer* (https://github.com/mrc-ide/MIPanalyzer) was used to assess parasite relatedness and detect population structure. We first performed principal component analysis (PCA) of all samples. Given the unwieldy size and lightly structured nature of the dataset, we subsetted samples by city for plotting, with samples coded by collection year. To more thoroughly determine population structure and relatedness, we calculated identity by descent (IBD) using maximum likelihood estimation. IBD networks connecting samples from each city with IBD ≥ 0.99 were generated using *ggraph*^68^ (version 2.1).

Complexity of Infection (COI), i.e. the number of parasite clones present in each sample, was calculated with *McCOILR* (https://ojwatson.github.io/McCOILR/index.html, version 1.3), an R implementation of *THE REAL McCOIL* (v2) categorical method^69^.

### Analysis of Putative Drug Resistance Genes and Potential Vaccine Candidates

All amino acid changes detected with the tiled gene panel and their respective frequencies were summarized using miplicoRn (https://github.com/bailey-lab/miplicorn, v0.2.1) and flextable^70^ (v0.9). We prioritized analysis of mutations that have previously been associated with drug resistance.

To detect potential selection on the potential vaccine candidates, we used vcf-kit (https://github.com/AndersenLab/VCF-kit) to calculate Tajima’s D in 100 bp sliding windows with a 50 bp step size.

Prior to CNV calculation, samples and MIPs were filtered for missingness as follows: samples with <5 unique molecular identifier (UMI) counts for more than 50% of MIPs were excluded, as were MIPs with <5 UMI counts for more than 50% of samples sequenced. For CNV calculation at loci of interest, the goal was to mitigate technical variations in the read coverage while maintaining true biological modifications. To correct the raw read data for systematic errors, z-score normalization of UMI coverage of each MIP was performed by accounting for potential sample and locus effects. Individual UMI coverages were transformed into z-score as 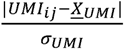, where *UMI_ij_* is the UMI coverage of a given MIP (or microhaplotype) *i* within a sample *j* and 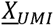 and *σ_UMI_* represent the mean of per sample means and standard deviations of UMI coverage, respectively. For each sample, the mean and standard deviation of UMI coverages were initially calculated across all loci included in the MIP panel. Consequently, this step generated distributions of means and standard deviations for the entire sample set in which we computed averages to obtain the parameters 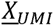 and *σ_UMI_*. The relative read abundance of a candidate gene within a sample, which corresponds to CNV, was estimated by dividing the z-score (*z_ij_*) of each MIP targeting this gene by the averaged z-score of all the other genes captured that are known not to be subject to CNV. As this method deals with sample and locus-derived errors and not batch effects, it was only applied to calculate CNV in samples that are sequenced together in the same run. The approach was previously validated in laboratory strains of *P. falciparum*, including 3D7, 7G8 and Dd2, in independent runs. Unlike 3D7 and 7G8, Dd2, the positive control with multiple copies of *mdr1*^71,72^ consistently showed CNV > 2 for this gene.

## Data Availability

Parasite sequence data is available through SRA (BioProject ID PRJNA1117913).

## Acknowledgements

We thank the study participants and various colleagues who have provided useful feedback on this project. Dr. Nicholas F. Brazeau, Dr. Valerie Gartner, Dr. Benjamin Redelings, and Sean V. Connelly provided assistance with computational techniques.

## Author contributions

ZRPH, JAB, JJJ, and HOV conceptualized and designed the study. JFS, DLP, and HOV supervised sample collection. ZRPH designed the MIP panels with input from KN, AAF, DJG, JAB, and JJJ using programs designed by ÖA. ZRPH, RC, AAF, and DJG performed laboratory analyses, data collection, and cleaning. ZRPH, KN, AS, and IK performed computational analyses. ZRPH, KN, AAF, JAB, JJJ, and HOV performed data analysis and interpretation. ZRPH, KN, JAB, JJJ, and HOV wrote the manuscript draft. All authors reviewed and approved the manuscript.

## Ethics

The protocol for this study was approved by the Institutional Review Board of the U.S Naval Medical Research Unit SOUTH (NAMRU SOUTH) in compliance with all applicable federal regulations governing the protection of human subjects (protocol NMRCD.2007.0004). Informed consent was obtained from all participants. For participants aged 8 to 17 years, written consent from parents or guardians and assent from participants were obtained.

## Disclaimer

The views expressed in this article are those of the author and do not necessarily reflect the official policy or position of the Department of the Navy, Department of Defense, nor the U.S. Government.

## Copyright

Some authors of this paper are employees of the U.S. Government. This work was prepared as part of their official duties. Title 17 U.S.C. §105 provides that ‘Copyright protection under this title is not available for any work of the United States Government.’ Title 17 U.S.C. §101 defines a U.S. Government work as a work prepared by a military service member or employee of the U.S. Government as part of that person’s official duties.

## Funding

This work was supported by the National Institute for Allergy and Infectious Diseases (R01AI165537, K24AI134990, and R01TW010870 to JJJ) and by the Armed Forces Health Surveillance Division (AFHSD), Global Emerging Infections Surveillance (GEIS) Branch, ProMIS ID P0091_24_N6.

**Correspondence** and requests for materials should be addressed to Zachary R. Popkin-Hall or Jonathan J. Juliano.

## Data availability

Parasite sequence data is available through SRA (BioProject ID PRJNA1117913).

## Code availability

Code used for analysis is available from GitHub at https://github.com/IDEELResearch

