## Supplemental Material for "High-Throughput Genotyping of *Plasmodium vivax* in the Peruvian Amazon via Molecular Inversion Probes"

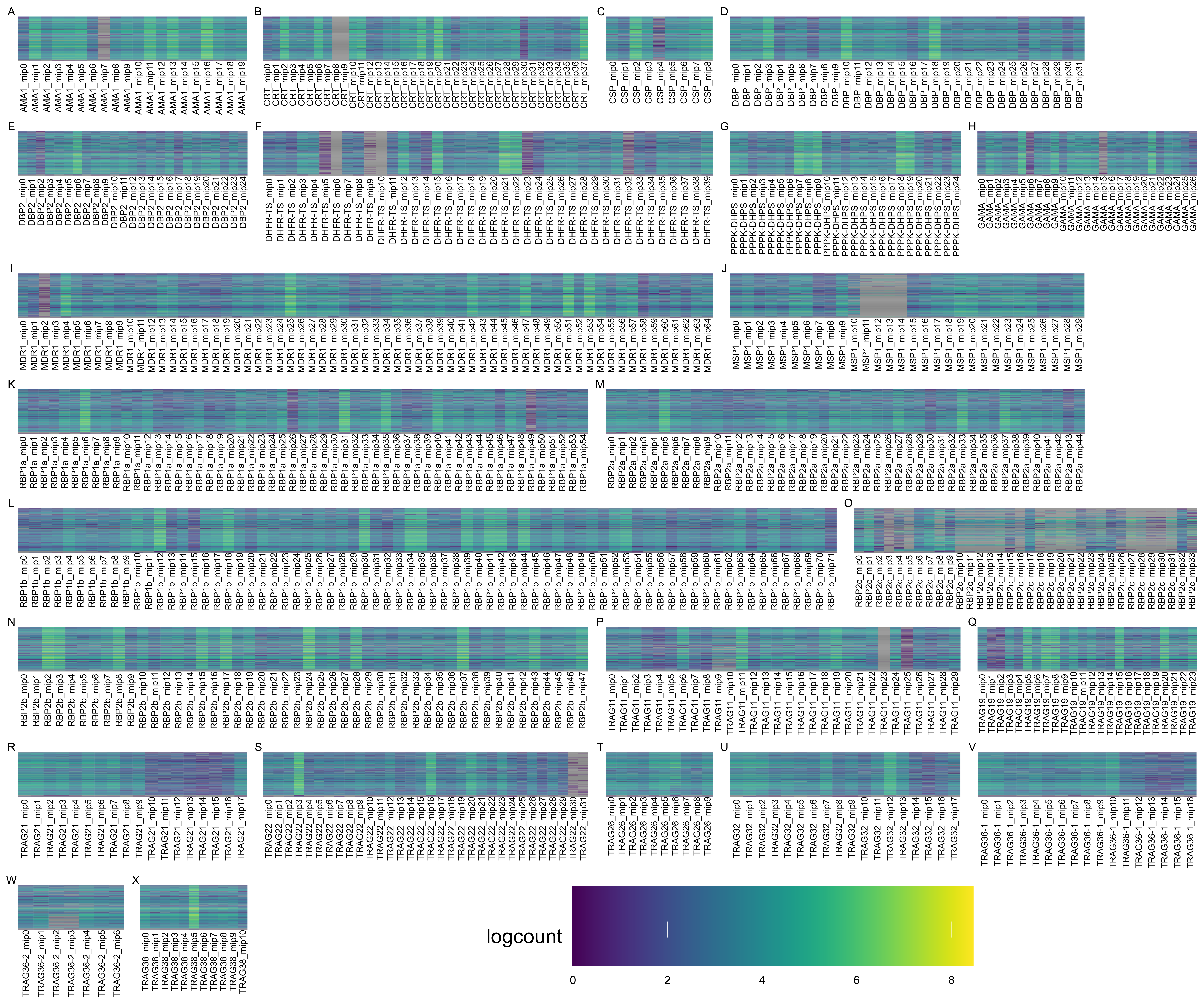

**Supplemental Figure 1 –** Heatmaps showing MIP sequencing performance for all tiled genes included in PvG panel. Sequencing performance is shown as log(UMI count) for each MIP.

**Supplemental Table 1 – Community and collection year of samples used in final analysis.**

| **Community** | **2012** | **2013** | **2014** | **2015** | **2016** | **2017** | **2018** | **Total** |
| --- | --- | --- | --- | --- | --- | --- | --- | --- |
| Alto Nanay |  |  |  |  | 1 | 1 |  | **2** |
| Belén | 31 | 133 | 41 | 9 | 8 | 47 |  | **269** |
| Indiana |  |  |  | 3 | 7 | 11 |  | **21** |
| Iquitos | 2 | 1 | 3 | 15 | 38 | 39 | 1 | **99** |
| Napo |  |  |  |  |  | 4 |  | **4** |
| Nauta |  |  |  |  |  |  | 1 | **1** |
| Pichanaqui |  |  |  |  | 1 |  |  | **1** |
| Punchana | 17 | 16 | 2 | 9 | 18 | 32 |  | **94** |
| Ramón Castilla |  |  |  |  | 1 | 2 |  | **3** |
| San Juan Bautista | 16 | 27 | 9 | 20 | 56 | 57 | 1 | **186** |
| Tigre |  |  |  |  | 1 |  |  | **1** |
| Torres Causana |  |  |  |  |  |  | 1 | **1** |
| **All Communities** | **66** | **177** | **55** | **56** | **131** | **196** | **4** | **685** |

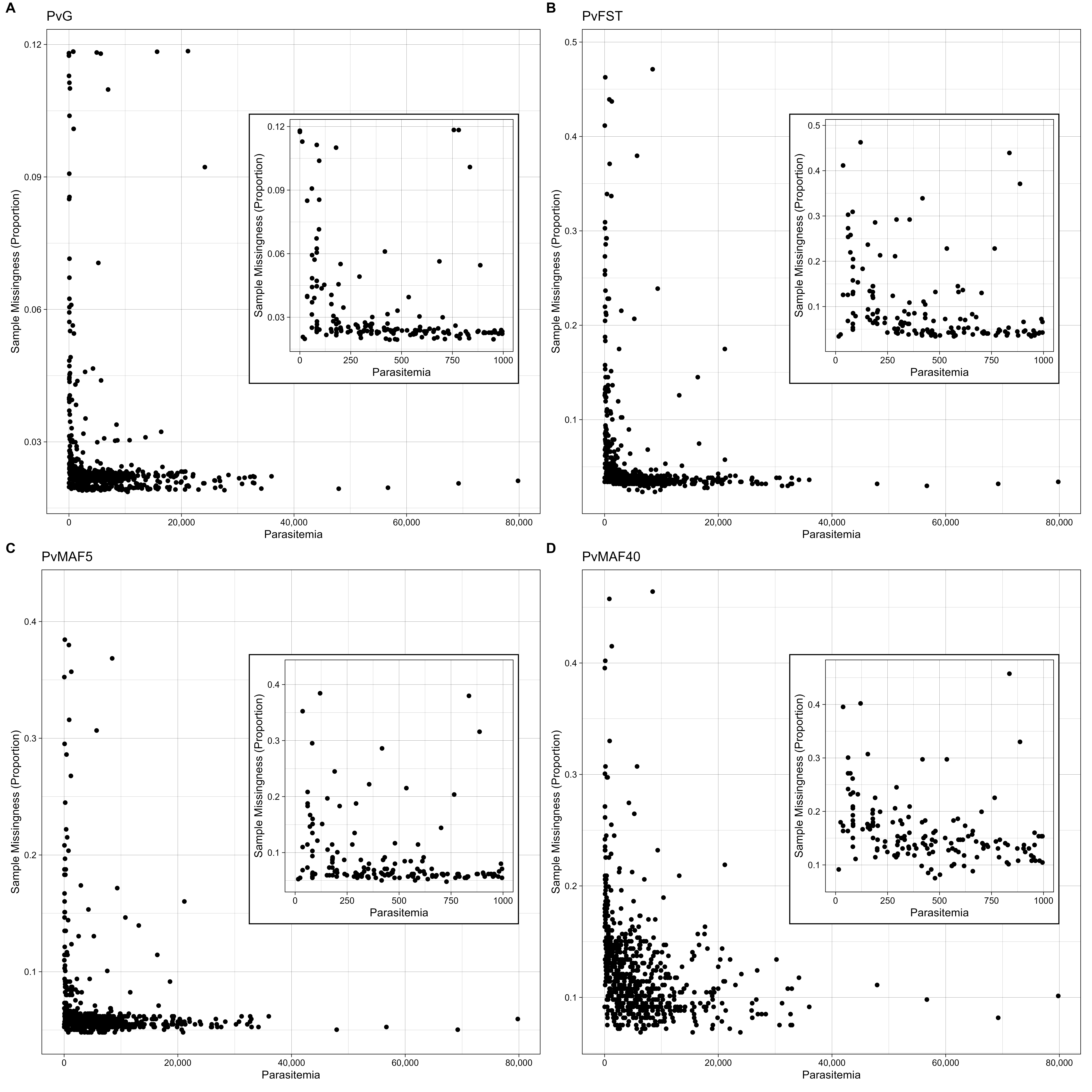

**Supplemental Figure 2** – Impact of asexual parasitemia (by qPCR) on sample missingness (proportion of sample not sequenced) for each MIP panel. Inset plots show values for samples under 1000 parasites/µL.

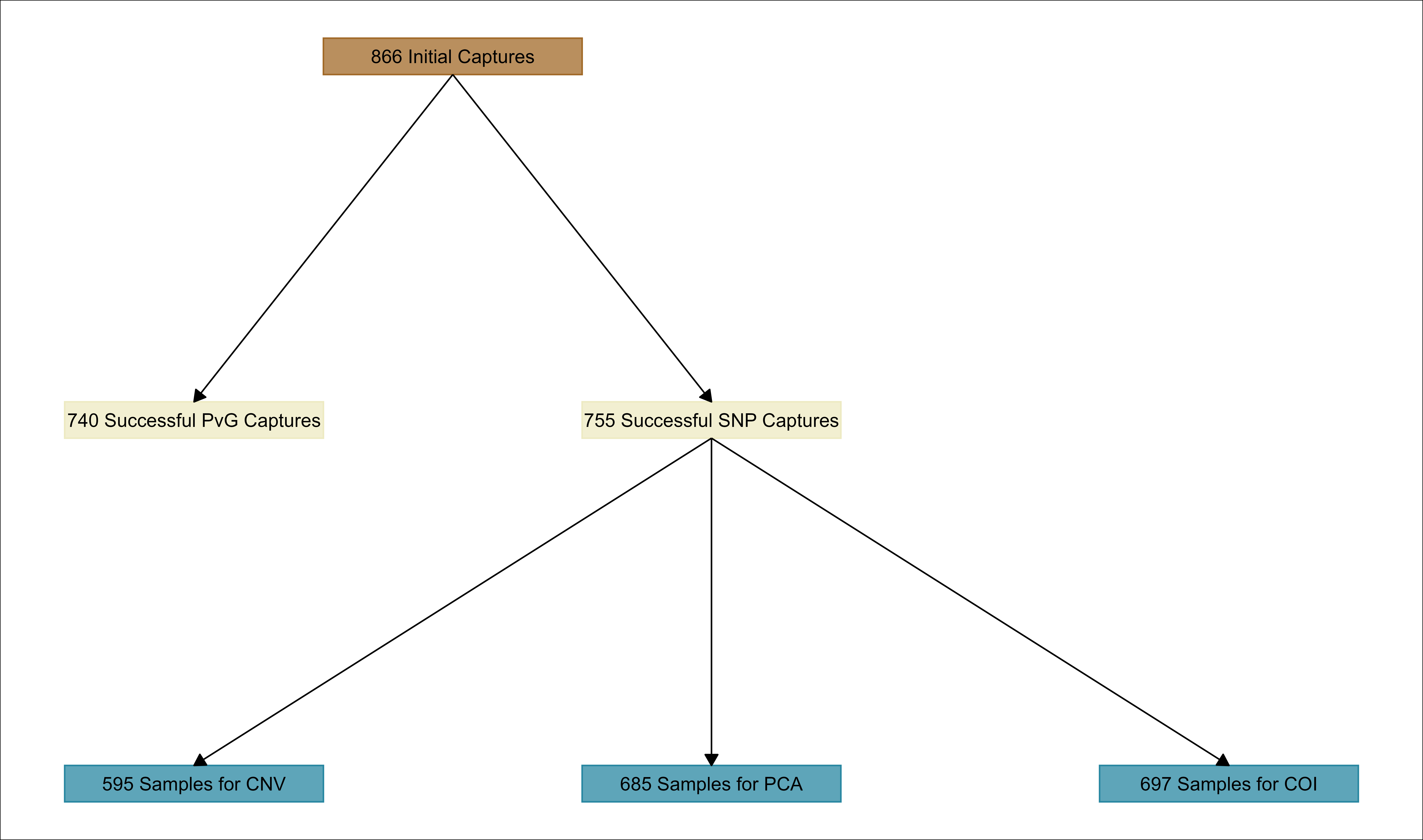

**Supplemental Figure 3** – Flowchart showing sample numbers retained for each analysis. SNP captures were only considered successful if samples sequenced for all three panels. Filtering requirements varied for copy number variation (CNV), principal component analysis (PCA), and complexity of infection (COI) analyses.

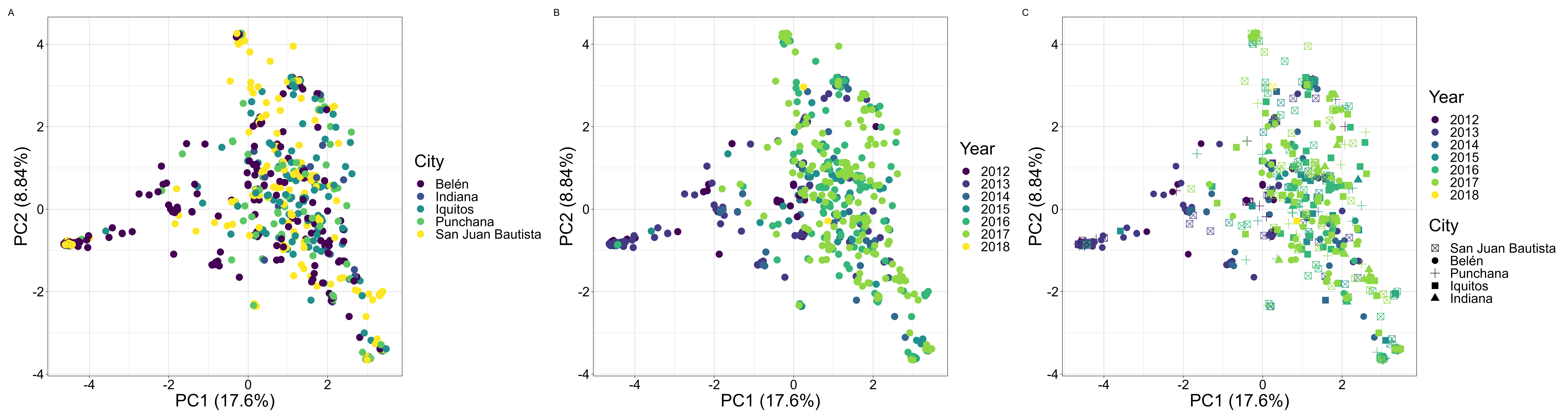

**Supplemental Figure 4** – Principal component analysis (PCA) plots based on SNP panels coded by A) city, B) year, and C) city and year.

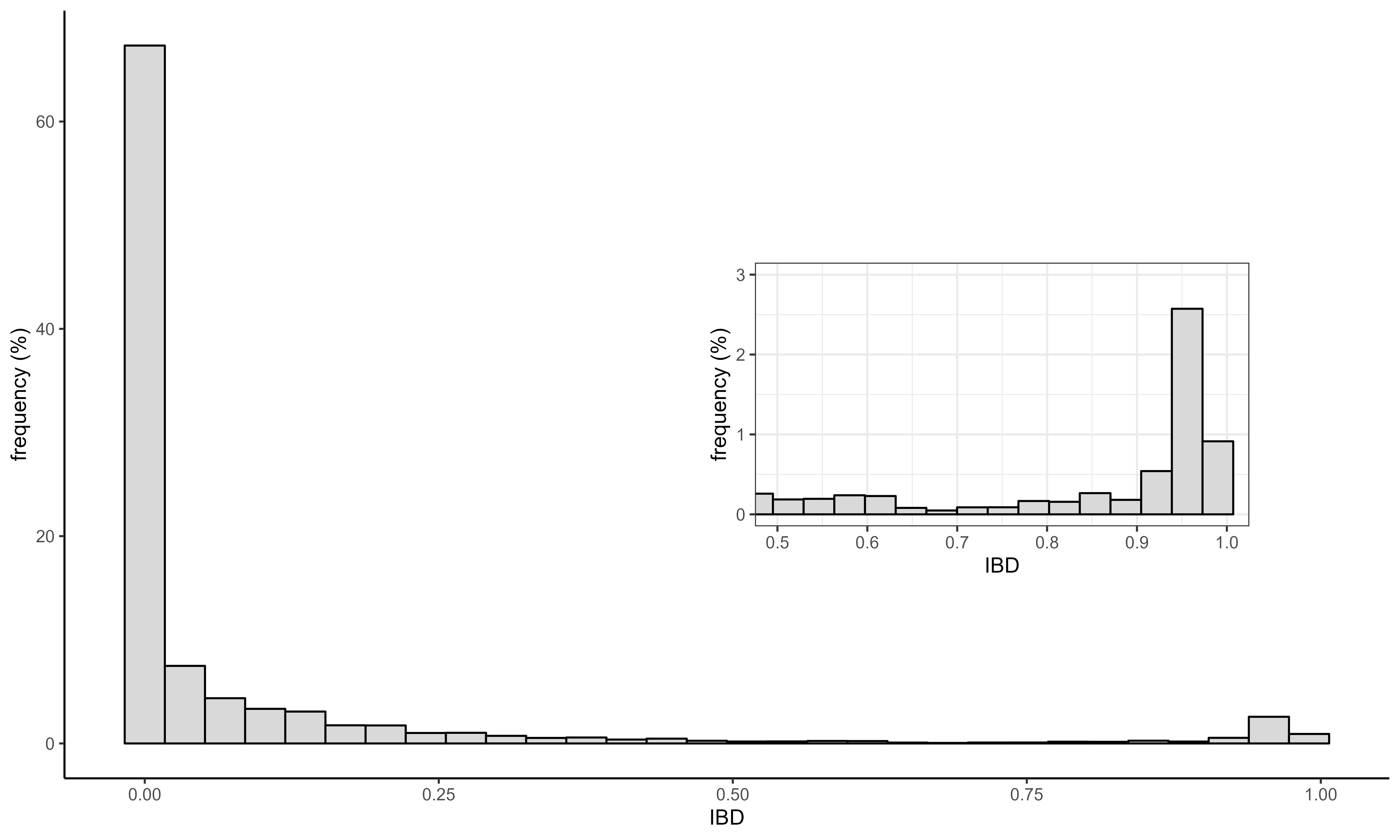

**Supplemental Figure 5** – Distribution of pairwise identity by descent (IBD) values for all samples included in PCA. Inset plot shows only those pairs with >0.5 IBD.

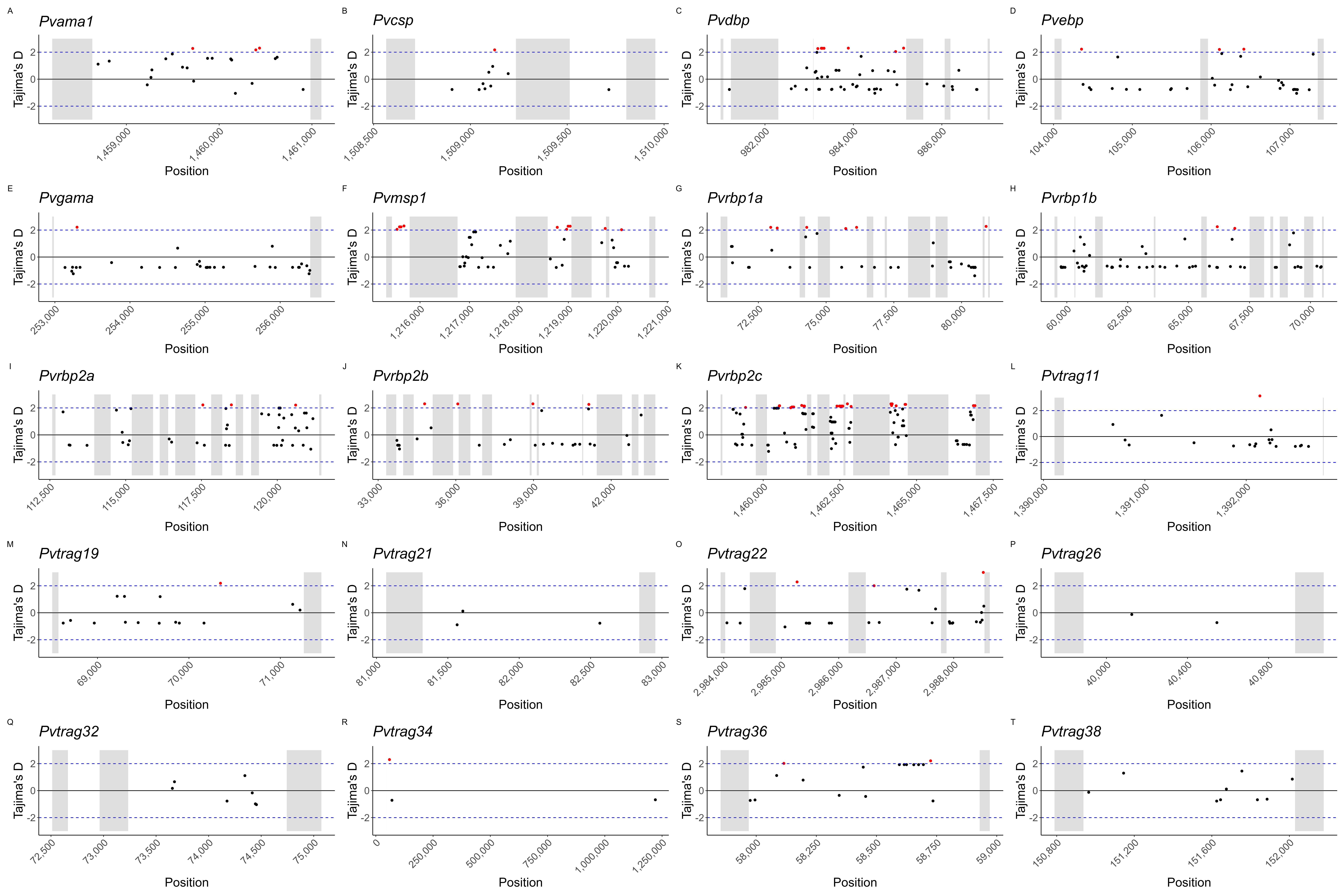

**Supplemental Figure 6** – Manhattan plots depicting Tajima’s D values in 100bp bins with a 50bp step size across potential vaccine target genes. Grey boxes indicate gene regions that were not captured by MIPs. Dots are placed in the midpoint of each bin, with Tajima’s D values >|2| highlighted in red.

**Supplemental Methods:**

*Global Variation in* Plasmodium vivax*:*

918 published *P. vivax* sequences representing all known NGS data were downloaded from the European Nucleotide Archive (accession numbers listed in **Supplemental Table 2**) using a custom bash script. Included in these 918 sequences were 12 lab strains^1,2^ and 17 samples from non-human animals^3,4^. The remaining samples are globally distributed across the endemic range of *P. vivax* and come from 27 countries (**Supplemental Table 3**)^1,5–17^. In addition to these published sequences, we also included sequences from collections in the Democratic Republic of the Congo and Peru.

**Supplemental Table 3 –** Samples included in global analysis of P. vivax variation for MIP design

| **Region** | **Country** | **Isolates** |
| --- | --- | --- |
| Africa | Eritrea | 12 |
| Africa | Ethiopia | 49 |
| Africa | Madagascar | 4 |
| Africa | Sudan | 5 |
| Africa | Uganda | 3 |
| Central America | Guatemala | 3 |
| Central America | Honduras | 28 |
| Central America | Mexico | 20 |
| East Asia | China | 7 |
| Europe | Spain | 1* |
| South America | Brazil | 51 |
| South America | Colombia | 34 |
| South America | Peru | 124 |
| South Asia | Afghanistan | 22 |
| South Asia | Bangladesh | 1 |
| South Asia | India | 44 |
| South Asia | Sri Lanka | 1 |
| South Asia | Pakistan | 32 |
| Southeast Asia | China | 4 |
| Southeast Asia | Cambodia | 184 |
| Southeast Asia | Indonesia | 8 |
| Southeast Asia | Laos | 2 |
| Southeast Asia | Myanmar | 28 |
| Southeast Asia | Papua New Guinea | 27 |
| Southeast Asia | Philippines | 1 |
| Southeast Asia | Thailand | 128 |
| Southeast Asia | Vietnam | 14 |

*Generated from ancient human cadaver, not modern isolate

Sequences were trimmed with cutadapt v1.18^18^, aligned to the PvP01 genome assembly^19^ using bwa-mem2 v2.2.1 ^20^, deduplicated with samblaster v0.1.24^21^, and filtered with samtools v1.14^22^ using a custom snakemake^23^ script. Quality control of aligned reads was performed using a custom snakemake script incorporating samtools and GATK v4.1.9.0^24^. Variant calling was performed in GVCF mode with GATK and bcftools v1.4^25^. Hypervariable regions^9^ were masked prior to variant filtering using vcftools v0.1.15^26^. Variant hard filtering was performed using GATK to exclude variants with a QD < 4, FS > 10, MQ < 50, MQRankSum < -2.5, or ReadPosRankSum < -5. These thresholds were identified by using the VariantsToTable tool within GATK to export metrics for all variants, the distributions of which were then plotted in R. Subsequently, bcftools was used to export only biallelic SNPs for further analysis.

Vcftools was used to calculate pairwise F_ST_ for all SNPs between all endemic regions (Southeast Asia/Papua New Guinea, South Asia, Africa, South America, and Central America) and the 400 SNPs with the highest F_ST_ (i.e. the best private alleles for each geographic region) were selected for MIP design along with previously identified SNPs of interest^5,27^. In addition to these differentiating SNPs, two sets of neutral (non-coding) SNPs were also generated. First, bedtools v2.30.0^28^ was used to export only non-coding SNPs from the original filtered VCF. Then, bcftools was used to generate one set of rare neutral SNPs (MAF < 0.05) and one set of common neutral SNPs (MAF > 0.4). These SNP sets were then further filtered to minimize the number of missing genotypes for included sequences: all included variants had a GQ > 90 and were genotyped in > 67% of individuals (MAF < 0.05) or > 55% of individuals (MAF > 0.4).

Lastly, we selected a set of genes associated with important cellular processes, such as cellular invasion or drug resistance, for design of tiled MIPs across the genome (**Supplemental Table 4**). Two broad categories of genes are of particular interest for *P. vivax* control and elimination: putative antimalarial resistance genes and red blood cell (RBC) invasion ligands that may present viable vaccine targets. While there are no experimentally validated antimalarial resistance markers in *P. vivax* due to the lack of an *in vitro* culture model, mutations in several genes have been associated with reduced antimalarial efficacy in field settings. These genes include chloroquine resistance transporter (*Pvcrt*), bifunctional dihydrofolate reductase-thymidylate synthase (*Pvdhfr-ts*), multidrug resistance protein 1 (*Pvmdr1*), and hydroxymethyldihydropterin pyrophosphokinase-dihydropteroate synthase (*Pvpppk-dhps*). Genes implicated in red blood cell invasion that present possible vaccine targets include apical membrane antigen 1 (*Pvama1*), circumsporozoite protein (*Pvcsp*), Duffy binding protein (*Pvdbp*), erythrocyte binding protein (*Pvebp*), GPI-anchored micronemal antigen (*Pvgama*), merozoite surface protein 1 (*Pvmsp1*), the reticulocyte binding proteins (RBPs, including *Pvrbp1a*, *Pvrbp1b*, *Pvrbp2a*, *Pvrbp2b*, and *Pvrbp2c*), and the tryptophan-rich antigens (TRAgs, including *Pvtrag11*, *Pvtrag19*, *Pvtrag21*, *Pvtrag22*, *Pvtrag26*, *Pvtrag32*, *Pvtrag34*, *Pvtrag36*, and *Pvtrag38*).

**Supplemental Table 4** – Genes targeted by PvG Panel

| **Gene Name** | **Gene ID (PvP01)** |
| --- | --- |
| AMA1 | PVP01_0934200 |
| CRT | PVP01_0109300 |
| CSP | PVP01_0835600 |
| DBP | PVP01_0623800 |
| DBP2 | PVP01_0102300 |
| DHFR-TS | PVP01_0526600 |
| GAMA | PVP01_0505600 |
| MDR1 | PVP01_1010900 |
| MSP1 | PVP01_0728900 |
| PPPK-DHPS | PVP01_1429500 |
| RBP1a | PVP01_0701200 |
| RBP1b | PVP01_0701100 |
| RBP2a | PVP01_1402400 |
| RBP2b | PVP01_0800700 |
| RBP2c | PVP01_0534300 |
| TRAG11 | PVP01_0532900 |
| TRAG19 | PVP01_1101400 |
| TRAG21 | PVP01_1401800 |
| TRAG22 | PVP01_1469800 |
| TRAG26 | PVP01_0000110 |
| TRAG32 | PVP01_0000170 |
| TRAG34 | PVP01_0700700 |
| TRAG36_1 | PVP01_0949200 |
| TRAG36_2 | PVP01_0000140 |
| TRAG38 | PVP01_0503600 |

*Pvcrt* has a well-supported role in chloroquine (CQ) resistance likely related to both mutations and variable expression levels^29^. Since CQ is still the first-line treatment for *P. vivax* in Peru and many other contexts^30^, monitoring putatively resistance-conferring mutations in this gene is critical to maintaining efficacious treatment programs. Previous studies have identified mutations in *Pvcrt* that were nearly at fixation^16^. However, since these mutations are intronic regions, the functional significance is unclear^16,29^. *Pvmdr1* is also putatively associated with CQ resistance^31^, as well as mefloquine (MFQ)^32^, amodiaquine (AQ)^33^, sulfadoxine-pyrimethamine (SP)^33^, and multiple other antimalarials^34^. Polymorphisms have been detected at heterogeneous levels throughout the Peruvian Amazon^31^, including in the CQ resistance-associated Y976F and F1076L mutations^31,35^.

Mutations in *Pvdhfr* and *Pvpppk-dhps* are associated with resistance to SP^36,37^, which is used as a partner drug in artemisinin combination therapy (ACT) for *P. falciparum*. In addition, while SP is not a preferred treatment for *P. vivax*, it is sometimes given in areas where *P. falciparum* and *P. vivax* co-occur, although it was removed as a first-line treatment in Peru in 2001^38^. As with *Pvcrt*, previous studies have identified mutations in *Pvdhfr* and *Pvpppk-dhps* ranging in frequency from very low (1.3%) to fixation in Peru^16,35,39^, which may have occurred while SP was still used for *P. falciparum* treatment in Peru^39^. Functional studies of these mutations have been limited to yeast and *E. coli* experimental models^40^.

*Pvama1* is a promising vaccine target due to its role in the invasion of multiple cell types, suggesting that it could function as a multi-stage vaccine^41^. *Pvama1* consists of four regions: 1) a pro-sequence, 2) a rich cysteine ectodomain, 3) a transmembrane domain, and 4) a cytoplasmic region^41^. The ectodomain contains three domains (DI, DII, and DIII)^41^. DI of the ectodomain has the highest mutation rate and highest genetic diversity^42^, while DII has high amino acid conservation and is the most immunogenic region^43^. Strong balancing selection has previously been identified in domain I of *Pvama1* with low diversity in the *Pvmsp1* C-terminal region in India^44^. While DII and DIII do not show evidence of balancing selection in India, Venezuela, or the China-Myanmar border^43,45–47^, there is evidence of positive selection on DII in Sri Lanka^48,49^, indicating that evolutionary pressures on this gene vary geographically. *Pvama1* haplotype also affects antibody responses^41^, so characterizing and monitoring mutations in this gene will support the effective deployment of such a vaccine.

*Pvcsp* is the ortholog of *Pfcsp*, which is the basis of the WHO-recommended RTS,S and R21 vaccines for *P. falciparum*^50,51^. CSPs are highly conserved within *Plasmodium* and are involved in sporozoite invasion of both the mosquito midgut and the liver^52^. *Pvcsp*’s utility as a potential vaccine target is complicated by the existence of two common strains, VK210 and VK247^53^. Both strains are present in the Amazon^49,54,55^, with high variability in Peruvian isolates^49^. *Pvcsp* consists of an N-terminal domain, central repeat region, and C-terminal domain, with the central repeat region likely being the portion of the gene that is most important for immunity^56^. Balancing selection has been detected in the central repeat region in Brazil^57^, while there is some evidence of directional selection in the N-terminal and C-terminal domains in multiple countries, especially in the VK210 strains^58^.

*Pvdbp* plays a critical role in RBC invasion in Duffy antigen/receptor for chemokines (DARC)-positive individuals, who make up the vast majority of people living in areas with high *P. vivax* transmission^59^. While there is growing evidence of *P. vivax* infecting DARC-negative individuals in African contexts^6,60,61^, a molecular mechanism for RBC invasion in this population has yet to be elucidated and *P. vivax* burdens in sub-Saharan Africa are much lower than they are elsewhere^62^. As such, and given the high frequency of DARC positivity in Latin American and Asian populations, PvDBP is a promising blood-stage vaccine candidate^59^. *Pvdbp* consists of five exons, the second of which encodes a large protein domain divided into six regions. Region II (RII) of *Pvdbp* encodes the Duffy-binding-like (DBL) domain, which is thought to play a major role in reticulocyte binding^59^. However, studies have demonstrated low *Pvdbp* immunogenicity in the Amazon region, and *Pvdbp* is highly polymorphic^59^. In addition, *Pvdbp* copy number variation (CNV) has been detected globally in isolates from both DARC-positive and DARC-negative individuals^8,63^. Previous studies in South America have identified variable selective regimes in *Pvdbp* RII, including diversifying selection^64^, positive selection^65^, and neutral evolution^66^, while studies in other endemic regions have identified balancing selection^66^.

*Pvebp*, formerly known as *Pvdbp2*, is a paralog of *Pvdbp*, but its role in RBC invasion is not clear. However, it may play a role in DARC-negative reticulocyte invasion and therefore may provoke natural immune response, thus serving as another viable blood-stage vaccine candidate^67^. *Pvebp* also exhibits CNV and strong positive diversifying selection, possibly in response to human immune response^67^.

*Pvgama* also plays a role in reticulocyte binding and is another blood-stage vaccine candidate^68^, but its population genetics have not previously been extensively studied.

*Pvmsp1* is a highly expressed gene on the merozoite surface, which along with its highly conserved nature, make it a leading vaccine candidate^44,55^.

The *Pvrbp* gene family plays an important role in invading reticulocytes^69–71^, are immunogenic, and are another potential vaccine target. While there are multiple functional *Pvrbp* genes, there are also partial and pseudogenes^69^. The *Pvrbp*s vary in their degree of polymorphism, e.g. *Pvrbp2c* is more polymorphic than *Pvrbp1*^72^, and their different domains may be under differential selective pressure^72^. CNV has previously been identified in *Pvrbp2a* and *Pvrbp2b*, with duplications detected in two Thai isolates^72^. Previous analyses have not identified deviations from neutrality in the *Pvrbp*s^66,73^.

Tryptophan-rich antigens (TRAgs) are another potential blood-stage vaccine target and are significantly expanded in *P. vivax* and its close relatives^74^. Multiple TRAgs are expressed on the merozoite surface and they function in lipid binding, with *Pvtrag25* binding preferentially to human reticulocytes via its C-terminal domain and the tryptophan/theonine-rich domain binding to sulfatide^74^. They may potentially serve as an alternate binding sites in DARC-negative individuals^74^, but their population genetics have not been analyzed in detail.

*MIP Design*:

Four MIP panels were designed using MIPTools (<https://github.com/bailey-lab/MIPTools>): PvFST (differentiating SNPs discussed above) PvG (tiled genes), PvMAF5 (rare non-coding SNPs), and PvMAF40 (common non-coding SNPs).

SNP panels were designed to be compatible with the Illumina NextSeq 2 x 75 bp sequencing run. This was accomplished by modifying the design settings file to have a maximum read length of 75 bp, a minimum read overlap of 10 bp, and a maximum read overlap of 20 bp. The PvG panel was designed to be compatible with the Illumina NextSeq 2 x 150 bp sequencing run. In this case, the design settings were less stringent, with a minimum overlap of 20 bp and a maximum overlap of 40 bp.

**Supplemental Results and Discussion:**

PvG capture and sequencing yielded 30,421,955 total UMI counts from 740 samples, 26 (3.5%) of which were flagged for uneven coverage. A total of 14,323 variants were identified after filtering based on within-sample allele frequency and UMI counts.

The SNP panels yielded between 29,429,868 and 69,527,709 on-target UMIs from 750 to 763 samples, with between 2 (0.27%) and 18 (2.4%) being flagged for uneven coverage. A total of 2,247 targeted variants were detected after filtering.

Sample missingness was very low (<12%) for PvG even at low parasitemia (≤100 parasites/µL). For the SNP panels, low parasitemia was a much larger impediment to successful capture and sequencing, although the median sample missingness was below 25% for all three panels.

Within *Pvama1*, a potential multi-stage vaccine target ^41^, we identified potential balancing selection in all three domains of the ectodomain. Strong balancing selection has previously been identified in domain I of *Pvama1* with low diversity in the *Pvmsp1* C-terminal region in India^44^. While domain II (DII) and DIII do not show evidence of balancing selection in India, Venezuela, or the China-Myanmar border^43,45–47^, there is evidence of positive selection on DII in Sri Lanka^48,49^, indicating that evolutionary pressures on this gene vary geographically. However, the presence of balancing selection throughout the gene in these Peruvian samples could present a problem for AMA1-based vaccines in South America.

Within *Pvdbp*, we found evidence of balancing selection in region II (RII), which encodes the Duffy-binding-like (DBL) domain, which is thought to play a major role in reticulocyte binding^59^, and has been determined to be subject to variable selective regimes in other South American studies, including diversifying selection^64^, positive selection^65^, and neutral evolution^66^, while studies in other endemic regions have identified balancing selection^66^. We also identified potential balancing selection in RIII, which encodes the Duffy-antigen binding domain, RVII, which encodes the erythrocyte binding antigen, and the cytoplasmic domain, regions of *Pvdbp* which have not been as thoroughly studied. As with AMA1, balancing selection in *Pvdbp* should be thoroughly evaluated prior to launching DBP-based vaccines.

Within *Pvebp*, we identified potential balancing selection in RII and RIII-V. *Pvebp*, formerly known as *Pvdbp2*, is a paralog of *Pvdbp*, but its role in RBC invasion is not clear. However, it may play a role in DARC-negative reticulocyte invasion and therefore may provoke natural immune response, thus serving as another viable blood-stage vaccine candidate^67^. However, both RII and RIII-V appear to have low antigenicity in Asian populations, where they are under positive selection and purifying selection, respectively^75^.

*Pvmsp1* is a highly expressed gene on the merozoite surface, which along with its highly conserved nature, make it a leading vaccine candidate^44,55^. However, we identified potential balancing selection in the C-terminal domain, in line with a study of Korean isolates that also identified balancing selection^76^.

We also identified evidence of balancing selection in *Pvrbp1a*, *Pvrbp1b*, *Pvrbp2a*, *Pvrbp2b*, and *Pvrbp2c*. Within *Pvrbp1a*, we detected balancing selection in RII, RIII, and RIV. A previous study that did not include South American samples identified purifying selection in RII, weak balancing selection in RIII, and potential selection in RIV, but low antigenicity in all of these regions^77^. In the China-Myanmar border region, *Pvrbp2a*, *Pvrbp2b*, and *Pvrbp2c* were all determined to be undergoing positive selection^78,^ while other analyses of the *Pvrbp*s have mostly not identified deviations from neutrality^66,73^.

Tryptophan-rich antigens (TRAgs) are another potential blood-stage vaccine target and are significantly expanded in *P. vivax* and its close relatives^74^. They may potentially serve as an alternate binding sites in DARC-negative individuals^74^, but their population genetics have not been analyzed in detail, so the implications of our detection of balancing selection are unclear.
